# An Interpretable Multimodal Framework for Student Mental Health Risk Assessment Using Temporal Embeddings and Fuzzy Inference

**DOI:** 10.64898/2026.05.16.26352630

**Authors:** Ayush Shah, Apurva Mehta, C.K. Bhensdadia

## Abstract

Mental health challenges among university students have increased due to academic pressure, lifestyle changes, and continuous digital engagement. Existing approaches for mental health assessment often rely either on self-reported psychological scales or isolated behavioral indicators, limiting their ability to capture complex temporal and contextual patterns.

This study proposes an interpretable multimodal framework for student mental health risk assessment using behavioral sensing, academic information, ecological momentary assessments (EMA), and psychometric survey data.

A bidirectional Long Short-Term Memory autoencoder is employed to learn latent temporal representations from day-level behavioral sequences, while graph embeddings capture structural relationships among students using similarity-based neighborhood graphs. These representations are fused with academic and survey-derived features and reduced using Principal Component Analysis and Uniform Manifold Approximation and Projection. K-means clustering is then applied to identify behaviorally distinct student groups.

Experimental analysis on the StudentLife dataset demonstrates meaningful clustering performance with a Silhouette Score of 0.4209 and Adjusted Rand Index stability of 0.6869. The identified clusters correspond to low-risk, moderate-risk, and high-risk behavioral profiles.

To improve interpretability and practical usability, a fuzzy inference system is introduced to compute mental risk, academic risk, and wellbeing indices using psychometric indicators including PHQ-9, PSS, PANAS, VR-12, and Big Five personality traits.

The results demonstrate the potential of combining multimodal behavioral modeling with interpretable fuzzy reasoning to support early mental health risk assessment in educational settings.

## 1 Introduction

Mental health among college and university students has emerged as a major public health concern due to increasing academic pressure, social expectations, lifestyle changes, and continuous digital engagement [1]. Students frequently experience stress associated with academic workload, career uncertainty, social relationships, and personal expectations. In addition, changes in sleep patterns, reduced physical activity, and increased screen time have been shown to negatively influence overall wellbeing and psychological health [2]. Although awareness regarding student mental health has improved significantly in recent years, many existing approaches remain largely theoretical and are often difficult to translate into practical, real-world interventions [3]. Effective mental health assessment systems must therefore consider practical constraints such as stigma, limited accessibility to professional support, cultural factors, and time constraints faced by students. Developing scalable and data-driven assessment frameworks can support earlier identification of mental health risks and contribute to improved student wellbeing [4, 5].

Recent advances in mobile sensing, behavioral analytics, and machine learning have enabled the development of computational frameworks capable of continuously monitoring behavioral patterns in real-world environments. Smartphone-based sensing systems can capture various aspects of daily student behavior, including physical activity, sleep patterns, social interaction, and device usage. Such behavioral signals provide valuable insight into psychological and emotional states without requiring continuous manual reporting. Prior studies have demonstrated that multimodal sensing data can support prediction of stress, depression, academic performance, and social functioning among students [6, 7].

This focused on understanding the relationship between behavioral and academic factors associated with student wellbeing. While the preliminary model provided useful insights into potential risk indicators, it relied on relatively simple statistical representations and lacked the ability to capture temporal behavioral dependencies and complex interrelationships between students.

The dataset used in this study consists of multimodal behavioral data collected from multiple heterogeneous sources, including smartphone sensing data, Ecological Momentary Assessment (EMA) responses, educational records, and psychometric survey data [6]. Each modality captures a distinct perspective of student behavior and wellbeing. Sensing data reflects behavioral patterns such as activity levels, conversation duration, and device usage. EMA responses provide self-reported emotional and psychological states collected during daily activities. Educational records capture academic engagement and performance, while survey responses provide information regarding personality traits, stress, wellbeing, and social connectedness.

A major challenge in computational mental health assessment lies in effectively integrating heterogeneous temporal and static data sources into a unified and interpretable framework. Time-series behavioral data contain sequential dependencies and evolving behavioral trends that require temporal modeling techniques. In contrast, academic and psychometric variables represent relatively stable static characteristics. Combining these modalities enables mental health assessment from multiple complementary perspectives rather than relying on isolated behavioral indicators [7].

To address this challenge, the proposed framework integrates temporal representation learning, graph-based modeling, clustering, and fuzzy inference within a unified multimodal architecture. Sequential behavioral data derived from sensing and EMA sources are modeled using a bidirectional Long Short-Term Memory autoencoder to capture temporal behavioral patterns over time. Static academic and psychometric features are combined with graph-based representations, where students are modeled as nodes and relationships are constructed based on feature similarity. Graph embeddings are then used to capture structural interactions and latent relationships among students. This multimodal representation enables the framework to identify complex behavioral patterns associated with varying levels of mental health risk.

The overall workflow of the proposed framework is illustrated in Figure 1. Experiment 1 focused on understanding and exploring the dataset along with building an initial stress prediction prototype using logistic regression. Experiment 2 focuses on temporal representation learning using time-series behavioral data and graph embeddings for unsupervised clustering of student behavioral profiles. Experiment 3 focuses on extracting interpretable psychometric features and integrating them into a fuzzy inference system capable of generating wellbeing indices and personalized recommendations.

**Figure 1:**
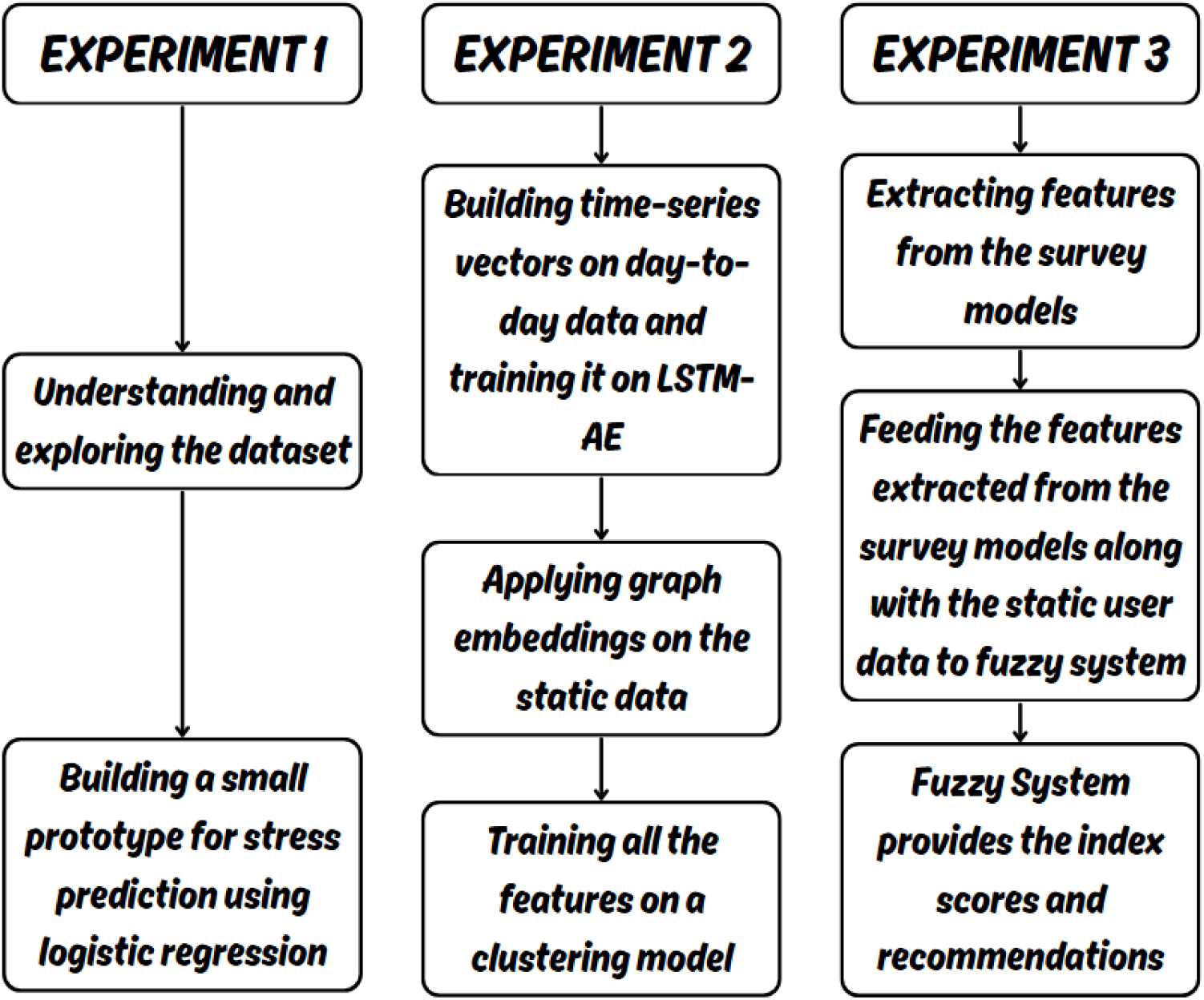
A brief overview on what is done and what is to be done

The primary objective of this study is to develop an interpretable and scalable framework for student mental health risk assessment using real-world multimodal behavioral data. Specifically, the proposed framework aims to:

- Integrate heterogeneous behavioral, academic, and psychometric data into a unified multimodal representation.
- Learn temporal behavioral patterns using deep sequential modeling techniques.
- Capture structural relationships between students using graph embeddings.
- Identify distinct student risk groups through unsupervised clustering.
- Develop an interpretable fuzzy inference system for generating mental risk, academic risk, and wellbeing indices.
- Provide actionable and personalized recommendations to support student wellbeing.

By combining machine learning, graph-based representation learning, and fuzzy reasoning, the proposed system aims to support early mental health risk assessment while maintaining interpretability and practical applicability in educational environments.

## 2 Related Work

Traditional approaches for mental health assessment primarily relied on structured clinical interviews, psychological questionnaires, and behavioral observations performed by psychiatrists and mental health professionals. Widely used assessment tools such as the Beck Depression Inventory (BDI), Hamilton Anxiety Rating Scale (HAM-A), and General Health Questionnaire (GHQ) were commonly employed to evaluate emotional and psychological wellbeing [8, 9, 10]. Although these methods remain clinically important, they are often time-consuming, difficult to scale, and dependent on subjective self-reporting and clinical accessibility [11]. Furthermore, the absence of automated behavioral monitoring limited the possibility of continuous and real-time mental health assessment [12].

Recent advances in machine learning, deep learning, and mobile sensing technologies have significantly transformed computational mental health research [13]. Smartphone-based sensing systems and wearable devices now enable continuous collection of behavioral signals such as mobility, activity patterns, social interaction, sleep behavior, and device usage. These developments have led to the emergence of data-driven frameworks capable of modeling psychological states using passive behavioral sensing.

Several studies have explored the use of passive smartphone sensing for stress and depression prediction. Bonafonte et al. investigated the contribution of different passively collected sensor modalities using neural network models including Fully Connected Networks (FCN) and LSTMs [14]. Their work utilized 125 behavioral features derived from WiFi, GPS, audio, and phone usage logs for stress classification and depression regression. The study demonstrated that multimodal sensing data can provide meaningful predictive signals for mental health assessment, although challenges related to feature imbalance, reproducibility, and model sensitivity remained unresolved.

The StudentLife framework introduced one of the earliest large-scale smartphone sensing studies focused specifically on student populations [6]. The framework continuously monitored behavioral signals including sleep, mobility, physical activity, conversation, and co-location, alongside Ecological Momentary Assessment (EMA) responses for stress and mood tracking. The study identified significant relationships between behavioral patterns and psychological outcomes, showing that increased academic workload corresponded to higher stress and reduced sleep. However, the framework primarily relied on correlation-based analysis and was limited by small sample size and declining EMA compliance over time.

Subsequent research extended these ideas using more advanced machine learning techniques. Rui Wang et al. combined passive smartphone sensing and EMA data from the StudentLife and CrossCheck studies to predict academic performance and depression-related outcomes [15]. Using Lasso regression and longitudinal statistical modeling techniques, the study achieved strong predictive performance for GPA and weekly depression status. Nevertheless, the work highlighted challenges associated with imbalanced datasets, longitudinal behavioral variability, and recall bias in self-reported assessments.

Deep learning approaches have also been explored for psychological state prediction. Mikelsons et al. proposed a fully connected neural network model for stress classification using GPS-derived mobility metrics and temporal variables [16]. Although the model achieved improved performance compared to baseline approaches, the study emphasized the need for richer behavioral representations and advanced feature learning techniques capable of capturing subtle temporal dependencies.

To address inter-subject variability in behavioral data, Shaw et al. introduced CALM-Net, a multitask learning framework based on LSTM autoencoders for personalized student stress prediction [17]. The model combined shared representation learning with personalized multilayer perceptrons to capture both global and individual behavioral characteristics. The framework demonstrated improved stress prediction performance on the StudentLife dataset; however, it required historical data for each individual student, limiting its applicability to entirely unseen users.

Beyond temporal modeling, graph-based learning and nonlinear dimensionality reduction techniques have recently gained attention for analyzing complex high-dimensional behavioral data. Delahoz-Domínguez et al. demonstrated the effectiveness of combining Uniform Manifold Approximation and Projection (UMAP) with K-means clustering for discovering meaningful structures in multidimensional datasets [18]. Their work showed that nonlinear manifold learning can preserve both local and global relationships while improving clustering interpretability and separation quality.

Interpretability has also become an important research direction in computational mental health systems. Liu et al. proposed the Adaptive Dynamic Attribute and Rule (ADAR) framework for scalable neuro-fuzzy inference in high-dimensional environments [19]. The framework introduced adaptive feature weighting and dynamic fuzzy rule management to improve both predictive performance and interpretability. Their results demonstrated that fuzzy reasoning systems can effectively balance transparency and modeling capability in complex behavioral applications.

In addition to student mental health assessment, mobile sensing has also been applied to broader psychiatric applications. Wang et al. investigated passive mobile sensing for assessing social functioning among patients with schizophrenia using multimodal behavioral features and tree-based ensemble learning methods [20]. Their findings demonstrated that behavioral sensing can support prediction of clinically relevant social functioning indicators, although performance was constrained by small sample sizes and limited generalizability.

Despite substantial progress in computational mental health research, several limitations remain across existing studies. Many frameworks rely on isolated sensing modalities, limited datasets, or purely predictive deep learning models with reduced interpretability. Other approaches focus primarily on correlation analysis or supervised prediction without integrating structural relationships between individuals. Furthermore, relatively few studies combine temporal behavioral modeling, graph-based representation learning, clustering, and interpretable reasoning within a unified multimodal framework.

To address these limitations, the proposed work integrates multimodal behavioral sensing, psychometric surveys, academic information, temporal representation learning, graph embeddings, unsupervised clustering, and fuzzy inference into a unified and interpretable architecture for student mental health risk assessment.

A comparative summary of representative studies, including their primary features, modeling approaches, and reported performance metrics, is presented in Table 1.

**Table 1:** Summary of representative studies related to computational mental health assessment.

| Paper | Features | Models | Performance |
| --- | --- | --- | --- |
| ADAR (Fuzzy) | High-dim adaptive features | Neuro-Fuzzy | Lower RMSE, fewer rules |
| UMAP + KMeans | Socio-economic indicators | UMAP + KMeans | High silhouette score |
| Passive Data | 125 features (WiFi, logs) | FCN, LSTM | RMSE: 3.5; F1: 61.4 |
| Social Sensing | 115 features, PCA | RF, XGB, ET | MAE: 2.17, $r=0.62$ |
| CALM-Net | Temporal + LSTM encoding | Multitask LSTM AE | F1: 0.594 |
| Phone Mental Health | Statistical features | Lasso, GBRT | Recall: 81.5%, MAE: 1.45 |
| Deep Psych Model | GPS + temporal | FCN | F1: 0.42 |
| StudentLife | Sleep, mobility, social | Correlation | $r$: $-0.38$ to $-0.52$ |

## 3 Materials and Methods

### 3.1 Dataset

This study utilizes the publicly available StudentLife dataset, which contains multimodal behavioral data collected from 48 university students over a continuous 10-week academic term [6]. The dataset combines passive smartphone sensing, Ecological Momentary Assessment (EMA) responses, academic information, and psychometric survey data, enabling comprehensive analysis of student behavioral and psychological patterns.

The sensing modality captures day-level behavioral information using smartphone-derived features such as conversation duration, screen usage, physical activity levels, audio characteristics, and phone inactivity periods. These behavioral indicators provide insight into mobility, social interaction, daily routines, and digital engagement.

EMA responses provide repeated self-reported measurements related to stress, mood, sleep quality, and emotional state collected throughout the academic term. Educational records include indicators associated with academic engagement and performance, while psychometric surveys provide standardized measures related to stress, wellbeing, personality traits, and social connectedness.

The psychometric modality includes the Patient Health Questionnaire (PHQ-9), Perceived Stress Scale (PSS), Positive and Negative Affect Schedule (PANAS), Flourishing Scale, Loneliness Scale, VR-12 Health Survey, and Big Five personality traits. Together, these modalities provide complementary behavioral, emotional, and psychological representations of student wellbeing.

Figure 2 illustrates the overall structure of the multimodal dataset used in this study.

**Figure 2:**
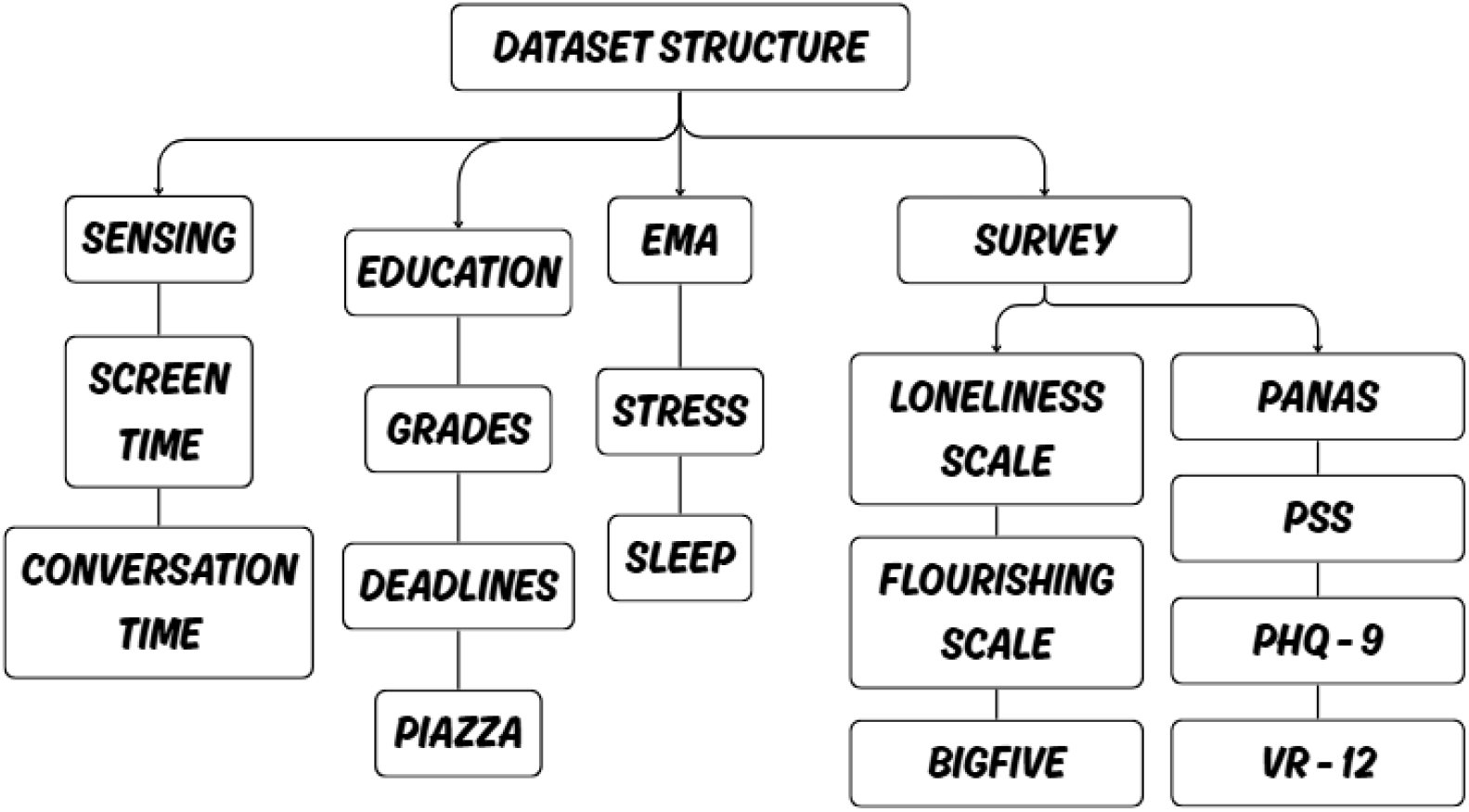
Structure of the StudentLife multimodal behavioral dataset consisting of sensing data, EMA responses, academic information, and psychometric survey data.

### 3.2 Data Preprocessing and Exploratory Analysis

The preprocessing pipeline was designed to standardize and integrate heterogeneous behavioral and psychometric data sources into a unified multimodal representation. Raw timestamps from sensing and EMA records were converted into consistent datetime representations and aggregated into day-level behavioral sequences for each student.

Missing EMA values were handled using mode-based imputation to preserve dominant behavioral trends while minimizing distributional distortion. Behavioral sensing features were subsequently normalized using RobustScaler to reduce the influence of extreme outliers and irregular behavioral fluctuations.

Static academic and psychometric variables were processed independently using StandardScaler normalization. Survey responses were transformed into normalized psychometric indices and derived feature representations. Time-series behavioral features and static survey features were then combined into a unified representation suitable for multimodal modeling.

Exploratory data analysis was additionally performed to examine relationships among behavioral, academic, and psychometric variables. Figure 3 presents a correlation heatmap illustrating relationships between selected behavioral and psychological indicators.

**Figure 3:**
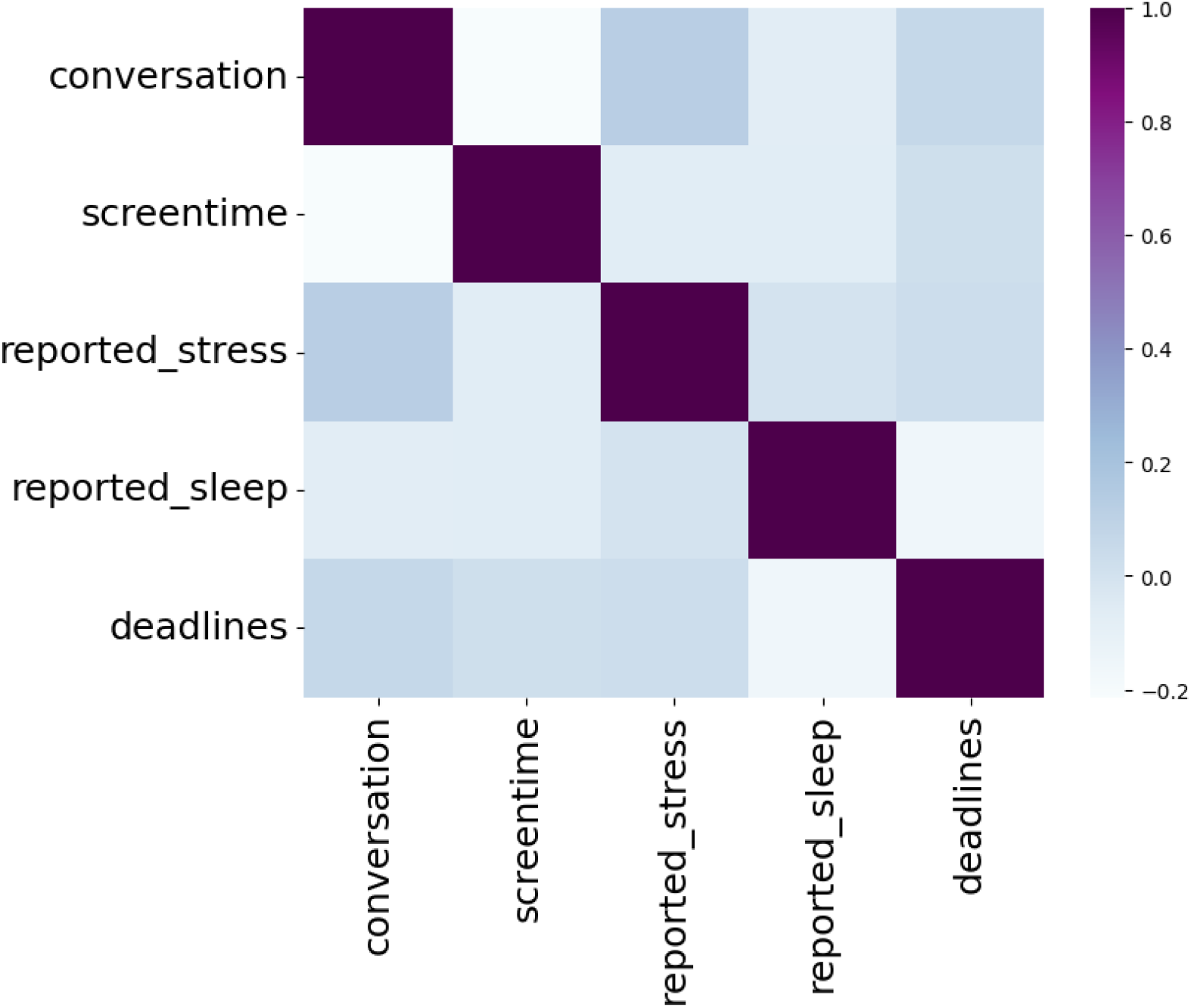
Correlation heatmap illustrating relationships among behavioral, academic, and self-reported psychometric features prior to multimodal fusion and representation learning.

To investigate the relationship between academic workload and stress, stress levels were compared across days with and without assignment deadlines. Figure 4 demonstrates increased average stress during academic deadline periods.

**Figure 4:**
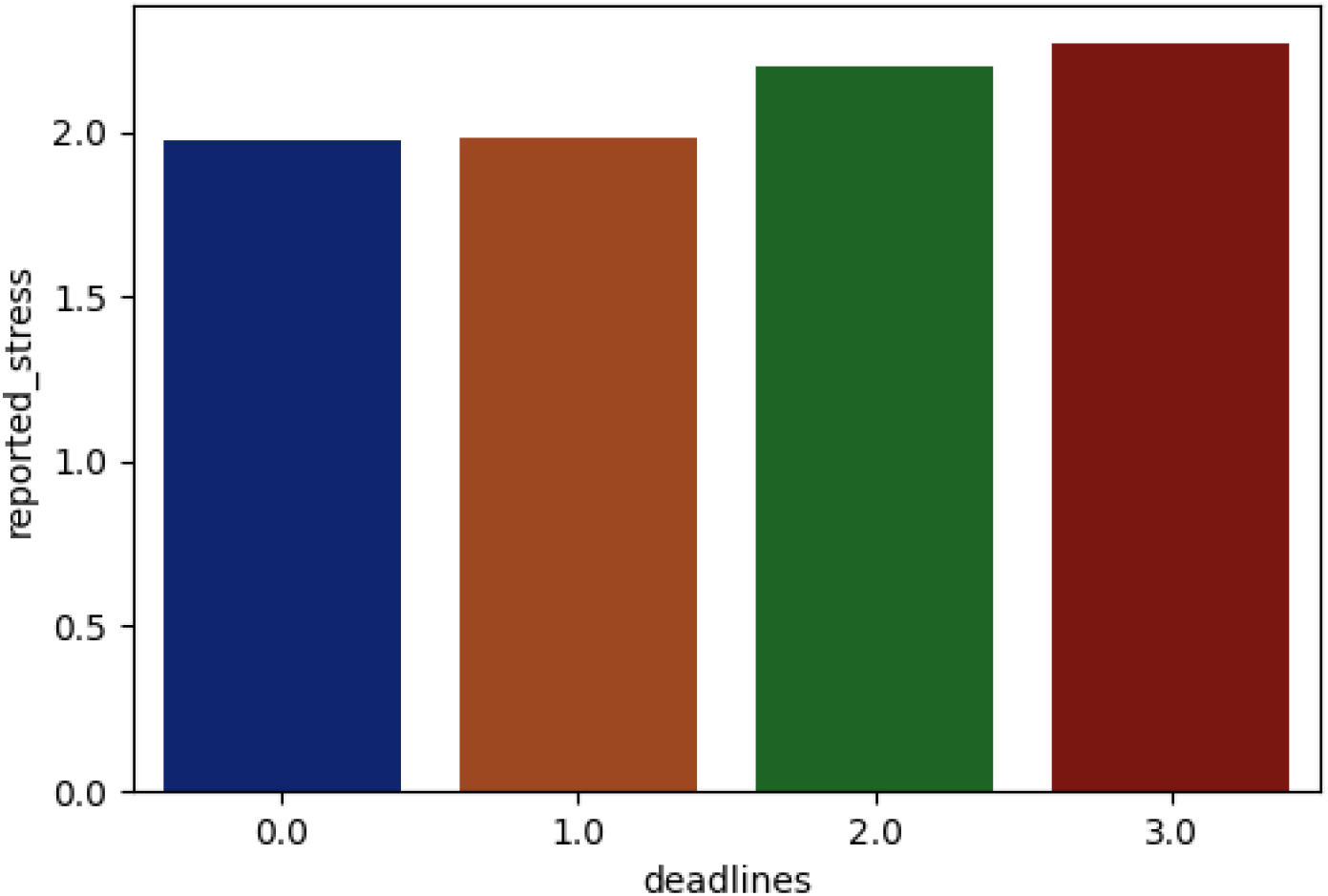
Comparison of average daily stress levels during assignment deadline and non-deadline periods.

### 3.3 Temporal Representation Learning

To model temporal behavioral dynamics, a bidirectional Long Short-Term Memory autoencoder (BiLSTM-AE) was implemented for sequential representation learning [21]. The architecture consists of an encoder-decoder framework designed to learn compressed latent representations from day-level behavioral sequences.

Bidirectional LSTM layers were employed to capture temporal dependencies in both forward and backward directions, enabling the model to learn evolving behavioral patterns across time. The encoder transforms sequential behavioral observations into low-dimensional latent embeddings, while the decoder reconstructs the original behavioral sequences from the learned representation.

The model was trained using the Adam optimizer with a learning rate of 0.0005 for 50 epochs. After training, encoder outputs were extracted as temporal behavioral embeddings representing student behavioral trajectories over time.

### 3.4 Graph-Based Representation Learning

To model latent structural relationships between students, graph-based representations were constructed using similarity relationships derived from static behavioral and psychometric features. A K-nearest-neighbor (KNN) graph was generated by connecting each student to the 15 most similar neighboring students based on feature similarity [22].

Similarity weights were computed using reciprocal distance weighting:

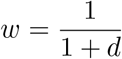

where *d* represents pairwise Euclidean distance between feature vectors. The normalized graph Laplacian was computed as:

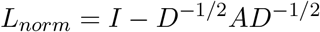

where *A* denotes the adjacency matrix and *D* represents the diagonal degree matrix. Spectral graph embeddings extracted from the normalized Laplacian were subsequently used to encode structural relationships among students [23].

### 3.5 Multimodal Feature Fusion and Dimensionality Reduction

Temporal embeddings learned from the bidirectional LSTM autoencoder were combined with graph embeddings and static psychometric features to generate a unified multimodal representation. This fusion strategy enables simultaneous modeling of temporal behavioral dynamics, structural student relationships, and stable psychological characteristics.

To reduce feature dimensionality and suppress noise, Principal Component Analysis (PCA) was applied prior to manifold learning [24]. Uniform Manifold Approximation and Projection (UMAP) was then employed to generate low-dimensional embeddings while preserving local neighborhood structure and nonlinear feature relationships.

The complete workflow of the proposed multimodal behavioral modeling framework is illustrated in Figure 5.

**Figure 5:**
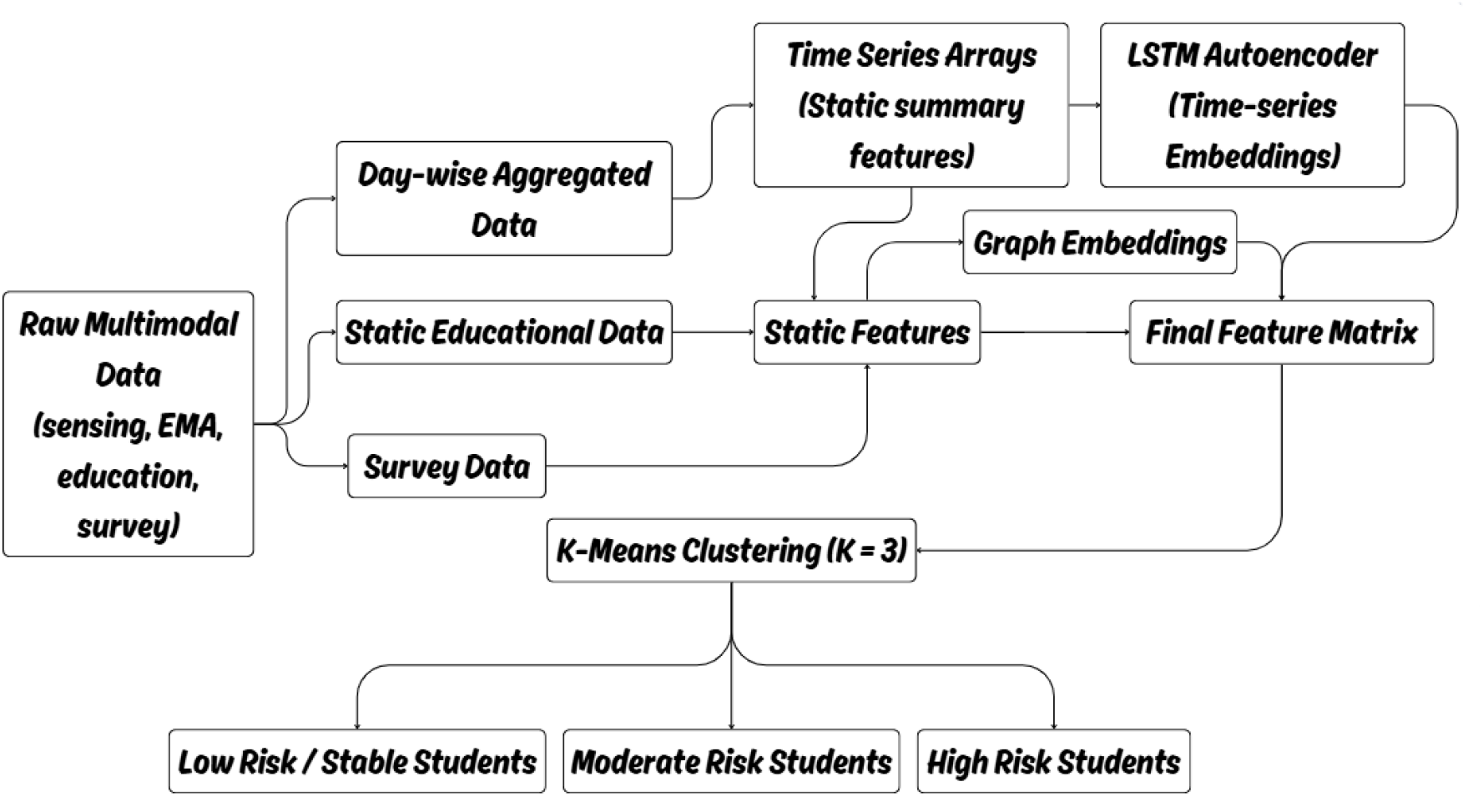
End-to-end workflow of the proposed multimodal framework integrating preprocessing, temporal representation learning, graph embeddings, multimodal fusion, dimensionality reduction, clustering, and fuzzy inference.

### 3.6 Clustering

Unsupervised clustering was performed on the reduced multimodal representations using the K-means algorithm with *k* = 3 clusters [18]. The clustering objective minimizes within-cluster variance according to:

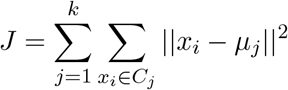

where *C*_*j*_ represents the *j*^*th*^ cluster and *µ*_*j*_ denotes its centroid.

The clustering process aimed to identify distinct behavioral risk groups associated with student mental health patterns. Cluster quality and stability were evaluated using Silhouette Score, Davies-Bouldin Index, Calinski-Harabasz Score, and Adjusted Rand Index.

The identified clusters corresponded to low-risk, moderate-risk, and high-risk student behavioral profiles. Table 2 summarizes the dominant behavioral characteristics associated with each cluster.

**Table 2:**
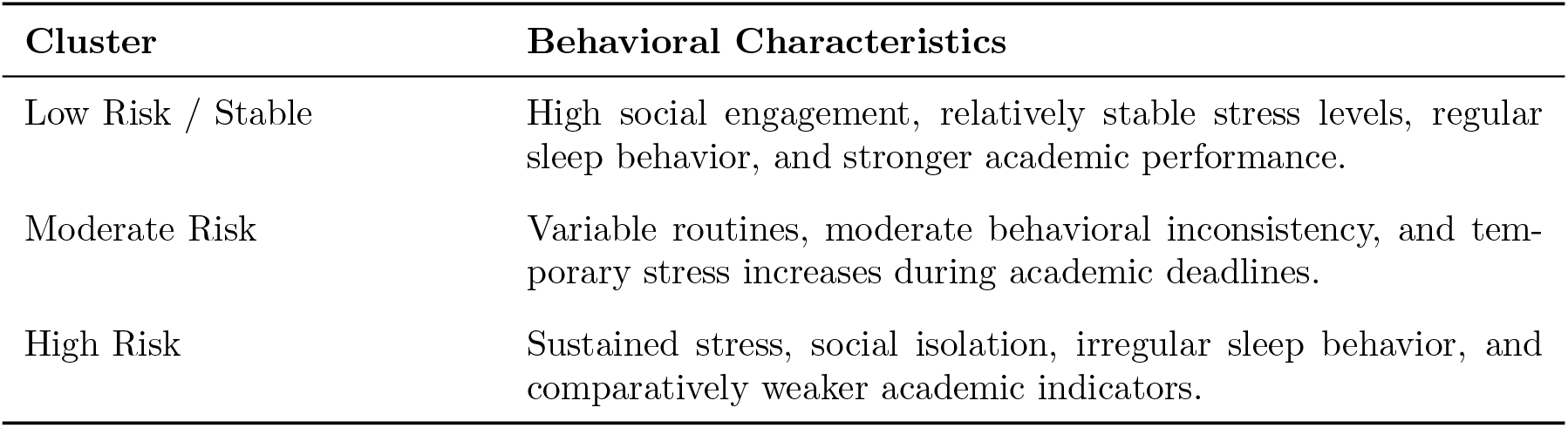
Behavioral characteristics associated with the identified student clusters.

### 3.7 Hybrid Multimodal and Fuzzy Inference Framework

To improve interpretability and practical usability, a hybrid multimodal and fuzzy inference framework was developed for behavioral risk assessment [19]. The framework combines psychometric survey modeling, behavioral indicators, and rule-based fuzzy reasoning to generate interpretable wellbeing measures.

#### 3.7.1 Psychometric Modeling

Separate computational models were developed for major psychometric surveys including PHQ-9, PSS, PANAS, Flourishing Scale, Loneliness Scale, VR-12, and Big Five personality traits. K-means clustering was used to extract representative psychometric patterns from survey responses.

The dataset can be represented as:

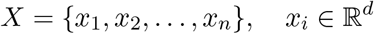

The K-means clustering objective minimizes within-cluster sum of squares:

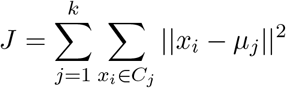

Cluster centroids were updated iteratively according to:

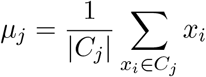

Outputs from these psychometric models were subsequently aggregated into a unified feature vector:

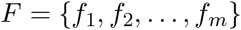

This feature vector was used as input to the fuzzy inference system.

#### 3.7.2 Fuzzy Inference System

A Mamdani-type fuzzy inference system (FIS) was implemented to compute:

- Mental Risk Index
- Academic Risk Index
- Wellbeing Index

Input variables were normalized to the range [0, 1] to ensure consistency across heterogeneous psychometric scales. Triangular membership functions were used to map normalized variables into low, medium, and high linguistic categories.

The triangular membership function is defined as:

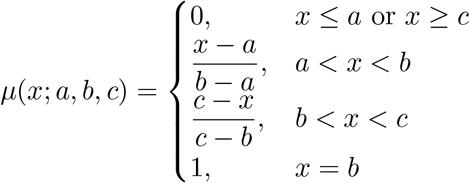

Rule-based inference was performed using min-max aggregation and weighted rule evaluation. Example fuzzy rules include:

IF depression is high AND stress is high THEN mental risk is high.

IF loneliness is high AND social interaction is low THEN wellbeing is low.

Three independent fuzzy engines were implemented for mental risk, academic risk, and wellbeing assessment.

#### 3.7.3 Recommendation Engine

A recommendation engine was developed on top of the fuzzy inference outputs to generate interpretable and actionable wellbeing suggestions. Recommendation rules were based on threshold-driven behavioral conditions derived from psychometric and behavioral risk scores.

Examples include:

- High stress → relaxation and stress-management recommendations
- High loneliness → social interaction recommendations
- Low academic indicators → structured study-planning recommendations

This module enhances practical applicability by transforming behavioral risk scores into understandable intervention-oriented guidance suitable for educational wellbeing applications.

#### 3.7.4 Prototype Deployment Interface

To demonstrate the practical applicability of the proposed framework, a lightweight prototype mobile application was developed for behavioral wellbeing assessment and personalized recommendation delivery. The application integrates the multimodal behavioral analysis pipeline and fuzzy inference framework within a user-oriented interface designed for continuous and accessible mental wellbeing monitoring.

The prototype application enables users to provide psychometric survey responses, behavioral indicators, and academic information through an interactive mobile interface. Input variables include responses from standardized assessment scales such as PHQ-9, PSS, PANAS, Flourishing Scale, Loneliness Scale, VR-12, and Big Five personality traits, along with behavioral attributes including screen time and academic performance indicators.

The collected information is processed through the proposed hybrid inference framework consisting of psychometric modeling, multimodal feature fusion, and fuzzy reasoning modules. Based on the computed behavioral representations and fuzzy risk scores, the application generates interpretable outputs including Mental Risk Index, Academic Risk Index, and Overall Wellbeing Index.

In addition to quantitative wellbeing scores, the system provides personalized recommendations derived from the rule-based fuzzy inference engine. These recommendations are intended to support behavioral awareness and early intervention by suggesting actionable strategies such as stress management techniques, improved sleep routines, social engagement activities, and structured academic planning.

Figure 6 presents the prototype mobile application interface. The assessment interface allows users to complete psychometric evaluations and provide behavioral inputs, while the results dashboard visualizes wellbeing indicators and personalized recommendations generated by the proposed framework.

**Figure 6:**
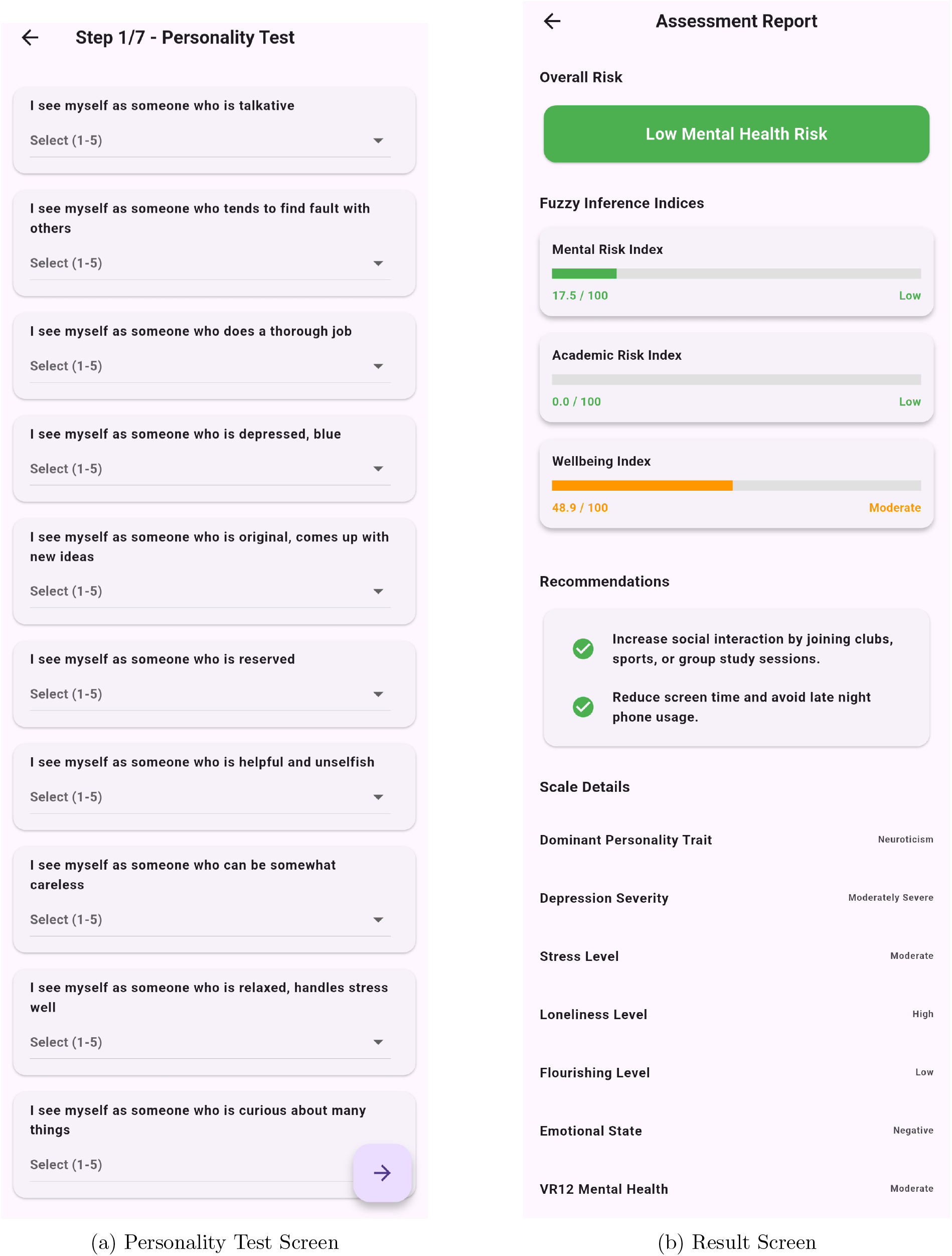
App Screenshots

The prototype demonstrates the feasibility of deploying the proposed behavioral assessment framework within real-world educational environments. By combining multimodal behavioral modeling with interpretable fuzzy reasoning in an accessible mobile interface, the system provides a foundation for future real-time student wellbeing monitoring and personalized intervention support.

## 4 Results

### 4.1 Clustering Performance Evaluation

The proposed multimodal framework demonstrated meaningful clustering performance across temporal, graph-based, and psychometric representations. The quality of the learned embedding space was evaluated using multiple clustering and representation-learning metrics including UMAP Trustworthiness, Silhouette Score, Calinski-Harabasz Index, Davies-Bouldin Index, and Adjusted Rand Index (ARI) stability.

The UMAP Trustworthiness score of 0.7491 Indicates that the nonlinear dimensionality reduction process preserved a substantial portion of local neighborhood relationships from the original high-dimensional feature space. This suggests that nearby behavioral representations in the original multimodal space remained structurally consistent after manifold projection.

K-means clustering achieved a Silhouette Score of 0.4209, indicating reasonably well-separated and interpretable behavioral clusters. Similarly, the Calinski-Harabasz Score of 53.1062 demonstrates meaningful inter-cluster separation and compactness, while the Davies-Bouldin Index of 0.7943 suggests favorable clustering quality with relatively low intra-cluster variance.

Cluster stability was additionally evaluated using repeated clustering experiments with Adjusted Rand Index (ARI) analysis. The mean ARI score of 0.6869 indicates relatively stable and reproducible cluster assignments across multiple runs.

A quantitative summary of the clustering and representation-learning performance is presented in Table 3.

**Table 3:** Quantitative evaluation of multimodal clustering and representation-learning performance.

| Metric | Value | Interpretation |
| --- | --- | --- |
| UMAP Trustworthiness | 0.7491 | Preservation of local neighborhood structure in the reduced representation space. |
| Silhouette Score | 0.4209 | Meaningful cluster separation and cohesion. |
| Calinski-Harabasz Score | 53.1062 | Well-separated and compact behavioral clusters. |
| Davies-Bouldin Index | 0.7943 | Low intra-cluster variance and improved cluster distinction. |
| Cluster Stability (Mean ARI) | 0.6869 | Consistent clustering behavior across repeated runs. |

Figure 7 illustrates the distribution of students across the identified behavioral clusters. The clusters appear relatively balanced, indicating that the multimodal representation learning framework avoided dominance by any single behavioral subgroup.

**Figure 7:**
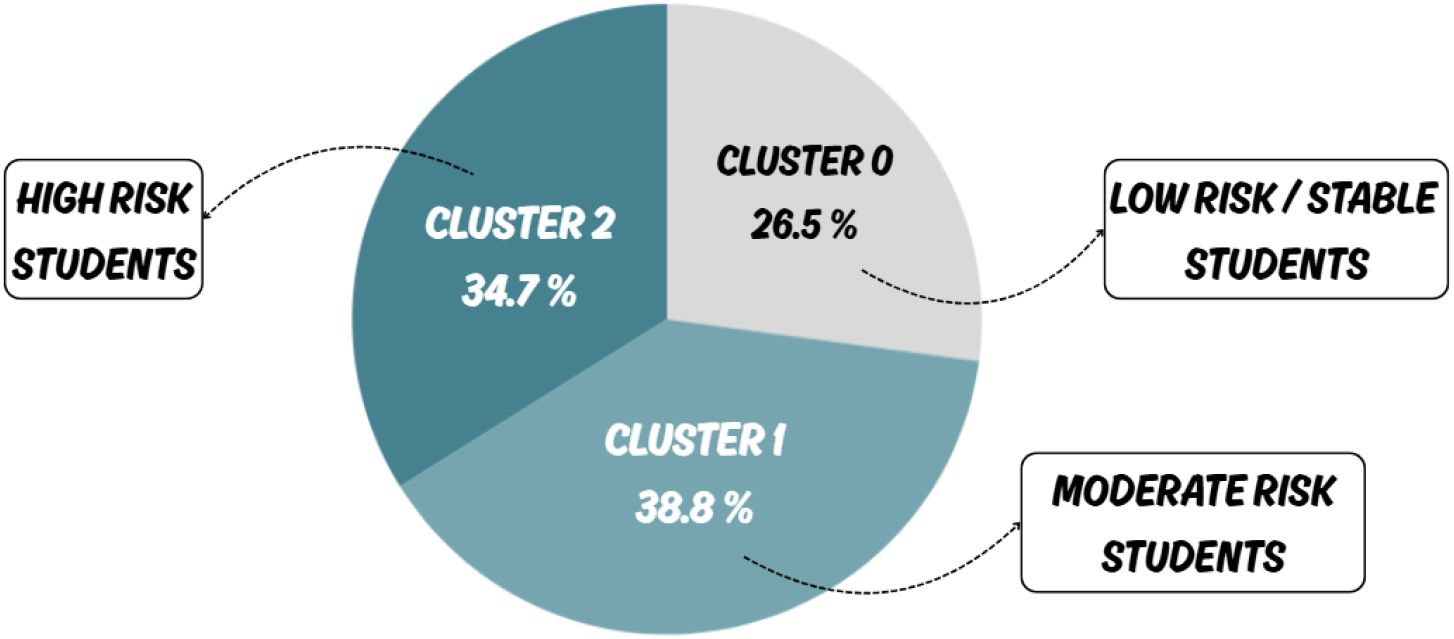
Distribution of students across the three behavioral clusters identified using multi-modal representation learning and K-means clustering.

### 4.2 Behavioral Cluster Analysis

The clustering framework identified three interpretable behavioral groups corresponding to low-risk, moderate-risk, and high-risk student wellbeing profiles. Distinct temporal and behavioral characteristics were observed across the identified clusters.

The low-risk cluster demonstrated relatively stable daily routines, stronger academic engagement, consistent sleep patterns, and higher social interaction. In contrast, the moderate-risk cluster exhibited fluctuating behavioral patterns and temporary increases in stress, particularly during periods associated with academic deadlines.

The high-risk cluster showed sustained stress patterns, reduced social interaction, irregular sleep behavior, and comparatively weaker academic indicators. These findings suggest that multimodal behavioral representations can effectively capture meaningful differences in student wellbeing profiles.

Table 4 summarizes the dominant behavioral characteristics associated with each identified cluster.

**Table 4:**
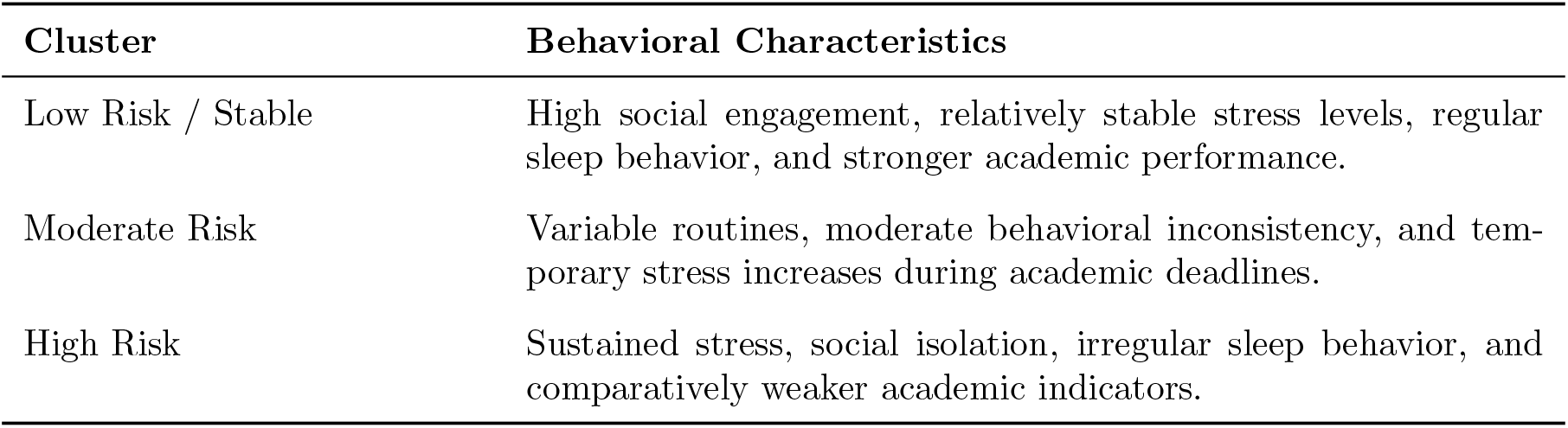
Behavioral characteristics associated with the identified student wellbeing clusters.

Figure 8 presents cluster-wise average temporal behavioral trajectories learned from the multimodal representation framework. The figure demonstrates clear behavioral separation between the three groups, indicating that temporal dynamics contributed substantially to cluster formation.

**Figure 8:**
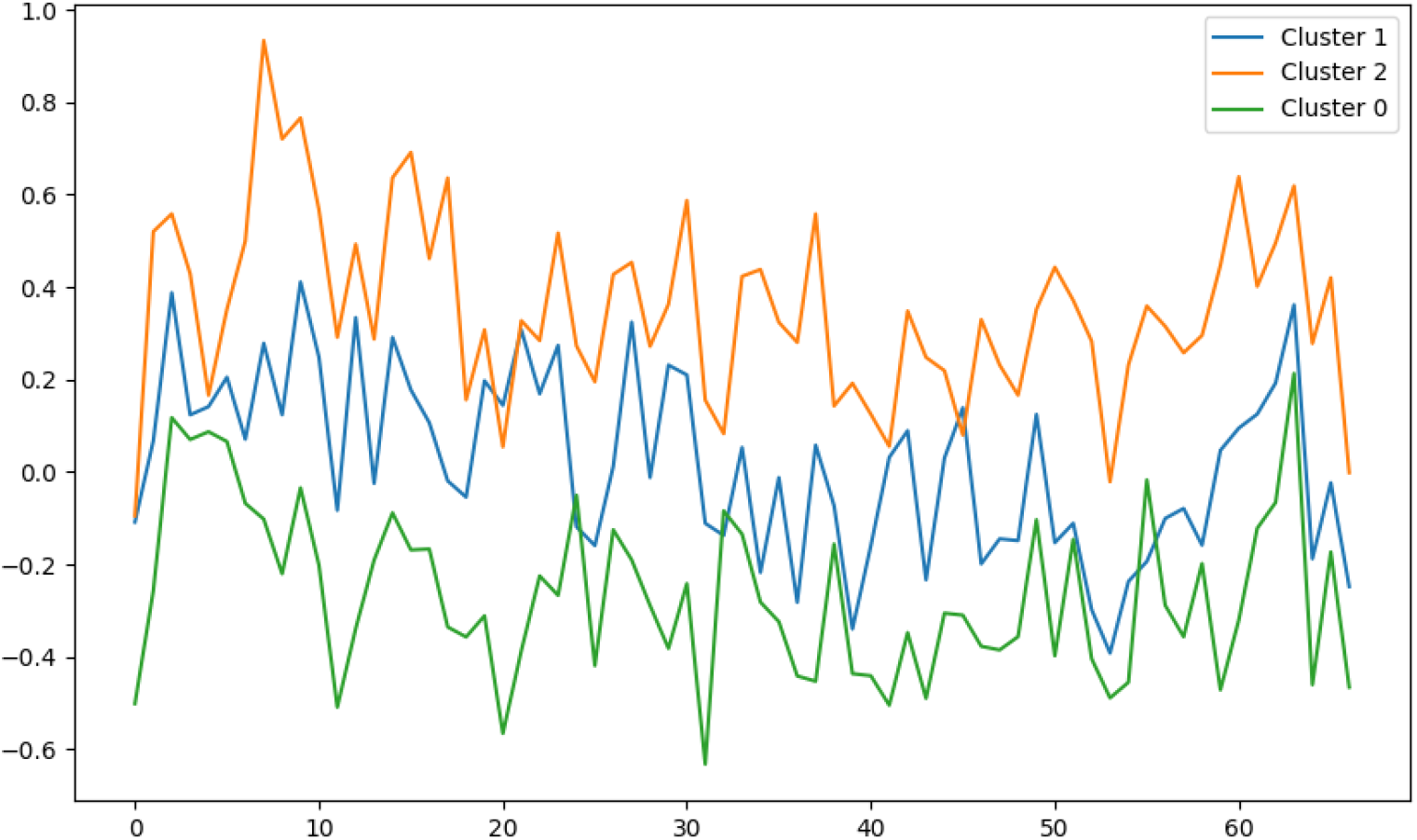
Cluster-wise average temporal behavioral patterns demonstrating separation between low-risk, moderate-risk, and high-risk student groups.

The observed separation between temporal behavioral trajectories indicates that sequential modeling captures meaningful behavioral evolution patterns associated with student wellbeing. In particular, the high-risk cluster exhibited sustained behavioral irregularities and elevated stress-related dynamics across time.

### 4.3 Temporal Pattern Analysis

To evaluate the effectiveness of temporal representation learning, reconstruction performance of the bidirectional LSTM autoencoder was analyzed using representative behavioral sequences.

The bidirectional architecture enabled the model to capture temporal dependencies in both forward and backward directions, improving sequential representation quality for variable-length behavioral sequences. The learned latent embeddings preserved dominant temporal characteristics while compressing behavioral information into lower-dimensional representations.

Figure 9 illustrates the reconstruction behavior of the bidirectional LSTM autoencoder for a representative student sequence.

**Figure 9:**
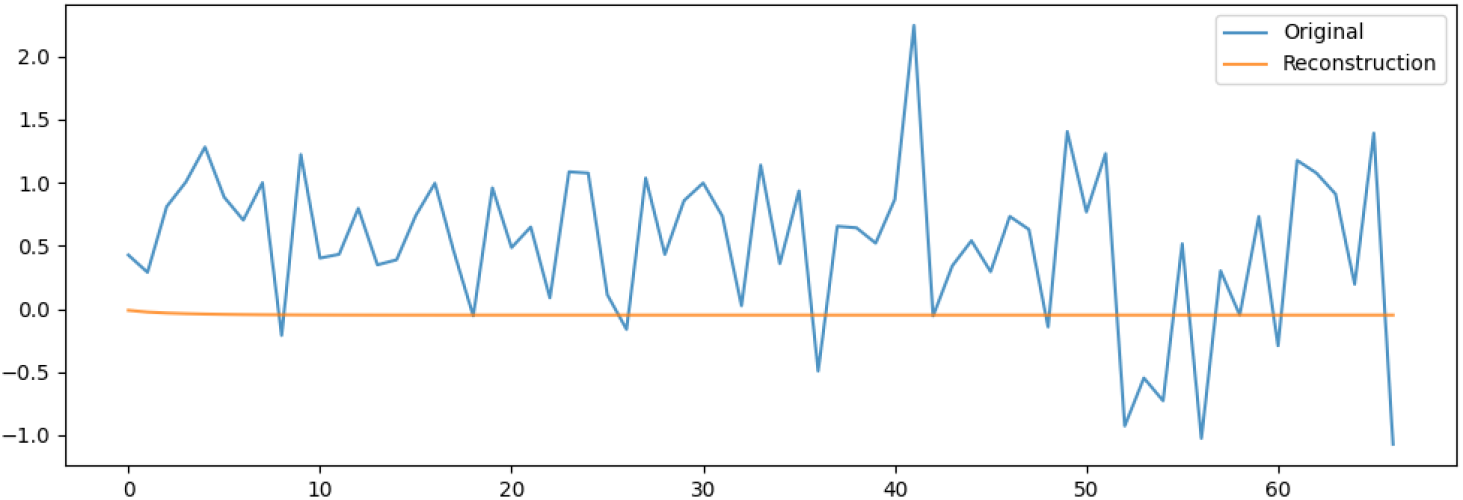
Reconstruction performance of the bidirectional LSTM autoencoder comparing original and reconstructed behavioral sequences for a representative student.

The reconstruction quality demonstrates that the proposed temporal modeling framework effectively captured dominant sequential behavioral characteristics while preserving long-term temporal dependencies. These temporal embeddings contributed significantly to the quality and interpretability of the final clustering structure.

### 4.4 Fuzzy Inference System Evaluation

The proposed hybrid multimodal and fuzzy inference framework generated interpretable wellbeing measures by integrating psychometric survey outputs, behavioral indicators, and academic features.

Separate psychometric modeling pipelines were constructed for PHQ-9, PSS, PANAS, Flourishing Scale, Loneliness Scale, VR-12, and Big Five personality traits. Clustering quality across these psychometric representations was evaluated using Silhouette Score and Davies-Bouldin Index.

The Big Five personality representation achieved the highest Silhouette Score of 0.62 and the lowest Davies-Bouldin Index of 0.54, indicating comparatively strong cluster separability. The Flourishing Scale and VR-12 representations also demonstrated relatively stable clustering structures.

Table 5 summarizes clustering quality across the psychometric assessment models.

**Table 5:** Comparative clustering quality across psychometric assessment models.

| Psychometric Model | Silhouette Score | Davies-Bouldin Index |
| --- | --- | --- |
| Flourishing Scale | 0.48 | 0.82 |
| Loneliness Scale | 0.21 | 1.67 |
| Big Five Personality Traits | 0.62 | 0.54 |
| Perceived Stress Scale (PSS) | 0.41 | 0.96 |
| VR-12 Physical Health Index | 0.55 | 0.71 |
| PHQ-9 Depression Scale | 0.37 | 1.08 |
| Static/Derived Features | 0.46 | 0.88 |

The fuzzy inference system subsequently integrated these psychometric representations into interpretable wellbeing scores including Mental Risk Index, Academic Risk Index, and Overall Wellbeing Index.

Figure 10 illustrates the workflow of the proposed hybrid multimodal and fuzzy inference framework.

**Figure 10:**
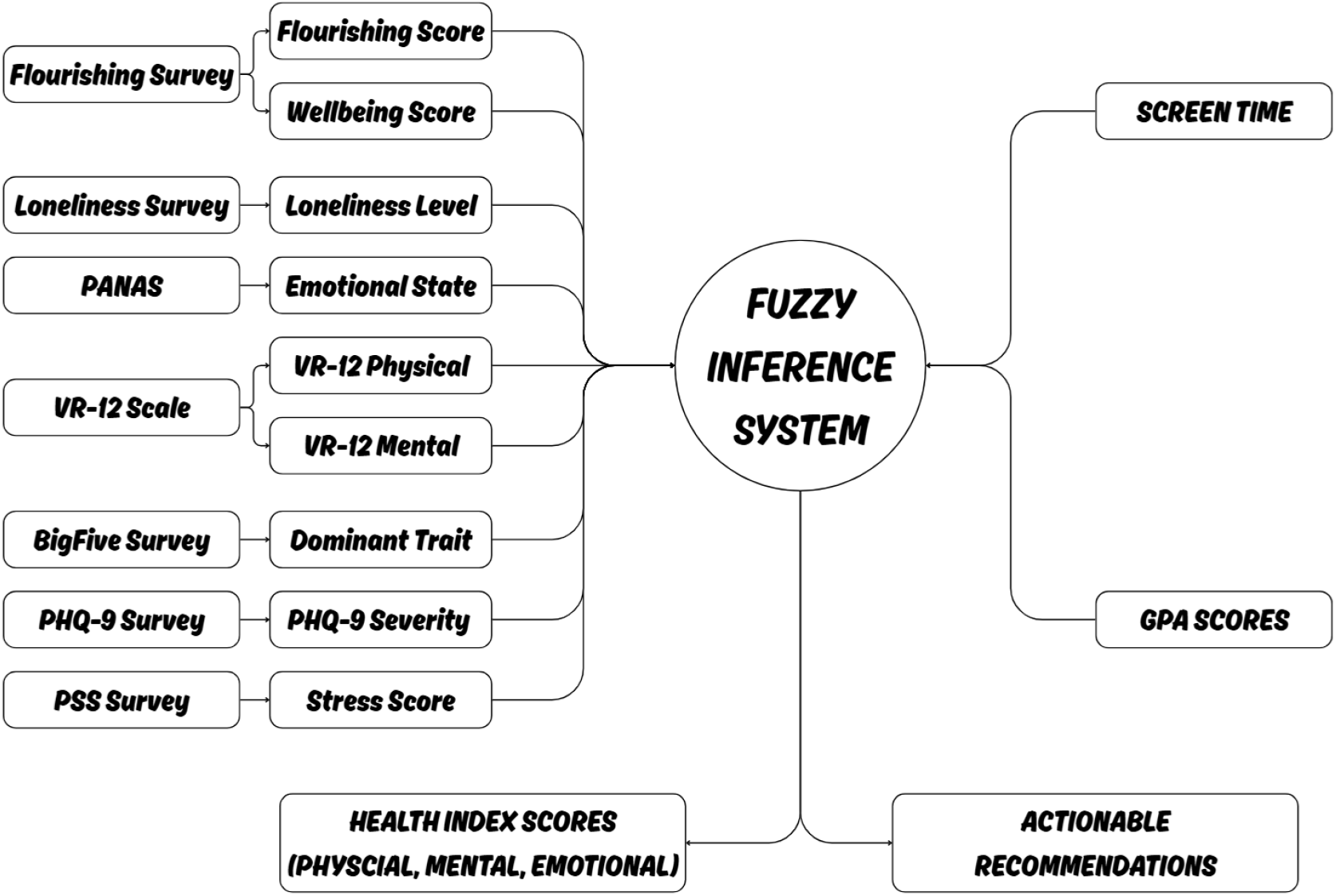
Workflow of the proposed hybrid multimodal and fuzzy inference framework for behavioral wellbeing assessment.

The fuzzy reasoning framework enabled interpretable mapping between behavioral indicators and wellbeing outcomes while preserving transparency in the decision-making process.

### 4.5 Qualitative Recommendation Analysis

In addition to quantitative wellbeing assessment, the proposed framework generated personalized behavioral recommendations using rule-based fuzzy inference. Recommendations were derived using threshold-based conditions associated with psychometric and behavioral risk indicators.

Examples of generated recommendations include stress-management guidance for students with elevated stress scores, social engagement suggestions for students exhibiting high loneliness indicators, and structured academic planning strategies for students with weaker academic behavioral patterns.

Table 6 presents representative behavioral patterns and corresponding recommendation outputs generated by the system.

**Table 6:** Representative recommendation outputs generated using the fuzzy inference framework.

| Behavioral Pattern | Generated Recommendation |
| --- | --- |
| High stress and irregular sleep | Stress management strategies and sleep routine stabilization guidance. |
| High loneliness and low social interaction | Social engagement and peer interaction recommendations. |
| Low academic engagement and increased screen time | Structured study planning and time-management recommendations. |
| Elevated depression indicators | Professional counseling awareness and wellbeing monitoring recommendations. |

The recommendation engine enhances the practical applicability of the proposed framework by transforming multimodal behavioral representations into interpretable and actionable wellbeing guidance suitable for real-world educational environments.

## 5 Discussion

The results demonstrate that multimodal behavioral fusion can effectively capture meaningful patterns associated with student mental health and wellbeing risk. By integrating temporal behavioral sequences, psychometric survey information, academic indicators, and graph-based structural relationships, the proposed framework provides a comprehensive representation of student behavioral dynamics in real-world educational environments.

The clustering results indicate the presence of distinct behavioral risk profiles among students. The identified low-risk, moderate-risk, and high-risk groups exhibited meaningful differences in stress patterns, sleep regularity, social interaction, and academic engagement. These findings are consistent with prior behavioral sensing studies demonstrating strong relationships between mental wellbeing, academic workload, sleep behavior, and social activity [6, 15].

The integration of bidirectional LSTM autoencoders enabled the framework to capture evolving behavioral trajectories rather than relying solely on static statistical summaries. Temporal modeling proved particularly useful for identifying sustained behavioral irregularities associated with elevated mental health risk. The reconstruction performance of the temporal model further demonstrated that the learned embeddings preserved dominant sequential characteristics while compressing high-dimensional behavioral information into interpretable latent representations.

Graph-based representation learning additionally improved the ability of the framework to model latent relationships among students. By constructing similarity-based neighborhood graphs, the system captured structural behavioral interactions that may not be identifiable through isolated individual-level analysis alone. The use of graph embeddings therefore enhanced the overall representational quality of the multimodal framework.

An important contribution of the proposed approach lies in its emphasis on interpretability. Many existing deep learning-based mental health prediction systems operate as black-box models with limited transparency regarding behavioral reasoning and decision generation. In contrast, the proposed fuzzy inference framework transforms multimodal behavioral representations into interpretable wellbeing indices and rule-based recommendations. This interpretability is particularly important in mental health applications, where transparent and explainable decision support systems are essential for practical adoption and ethical deployment.

The psychometric modeling component further demonstrated that different psychological assessment scales contribute distinct behavioral representations. Personality-related features, particularly those derived from the Big Five Inventory, showed relatively strong cluster separability, suggesting that stable psychological traits may influence broader behavioral patterns associated with wellbeing. Similarly, flourishing and stress-related measures demonstrated meaningful relationships with behavioral clustering outcomes.

The recommendation engine extends the practical applicability of the framework by converting behavioral risk indicators into actionable guidance. Rather than functioning purely as a predictive model, the system provides interpretable suggestions related to stress management, sleep stabilization, social engagement, and academic planning. Such behavioral intervention support may be valuable for proactive student wellbeing monitoring in educational institutions.

Compared with prior studies, the proposed framework provides several methodological advantages. Earlier works such as StudentLife primarily relied on correlation analysis and statistical modeling, while subsequent approaches focused largely on supervised prediction tasks using isolated sensing modalities [6, 15]. More recent deep learning approaches improved temporal modeling capability but often lacked interpretability or multimodal integration [17, 16]. The proposed work addresses these limitations by combining temporal representation learning, graph-based structural modeling, multimodal feature fusion, unsupervised clustering, and fuzzy reasoning within a unified framework.

Despite these promising results, the proposed framework should not be interpreted as a clinical diagnostic system. The identified clusters represent behavioral risk-oriented groupings rather than medically validated psychiatric diagnoses. Instead, the system is intended to function as an interpretable behavioral assessment and early-risk identification framework capable of supporting wellbeing awareness and intervention-oriented guidance.

Overall, the findings demonstrate the potential of combining multimodal behavioral sensing, representation learning, clustering, and fuzzy inference for scalable and interpretable student wellbeing assessment. The framework establishes a foundation for future real-time behavioral monitoring systems capable of supporting personalized mental health intervention strategies in educational settings.

## 6 Limitations

Several limitations should be considered when interpreting the findings and practical applicability of the proposed framework.

First, the study utilizes the StudentLife dataset, which consists of behavioral data collected from only 48 students within a single academic institution over a limited 10-week duration [6]. The relatively small sample size and restricted demographic diversity limit the generalizability of the findings across broader student populations, educational environments, and cultural contexts.

Second, a portion of the dataset relies on self-reported Ecological Momentary Assessment (EMA) responses and psychometric survey data. Although such assessments provide valuable psychological insight, self-reported measurements are inherently susceptible to recall bias, response inconsistency, and participant subjectivity. In addition, declining survey compliance over time may affect the reliability and temporal continuity of the collected data.

Third, the proposed framework performs behavioral risk stratification rather than clinical diagnosis. The identified low-risk, moderate-risk, and high-risk clusters represent computationally derived behavioral groupings based on multimodal sensing and psychometric patterns. These clusters should not be interpreted as medically validated psychiatric conditions or substitutes for professional clinical evaluation.

Fourth, the framework primarily focuses on unsupervised behavioral modeling and interpretability rather than clinically supervised prediction. Although the clustering and fuzzy inference components demonstrated meaningful behavioral separation and interpretable risk assessment, the absence of clinically annotated longitudinal outcomes limits direct evaluation of diagnostic accuracy and intervention effectiveness.

Fifth, the multimodal behavioral representations are dependent on smartphone-derived sensing data and digital behavioral indicators. Variations in device usage behavior, sensor availability, operating system restrictions, and participant interaction patterns may introduce inconsistencies across data collection environments. Furthermore, smartphone sensing alone may not fully capture broader environmental, social, or physiological factors associated with mental wellbeing.

Sixth, the fuzzy inference system relies on manually designed linguistic rules and threshold-based membership functions. Although this improves interpretability, the selection of membership ranges and inference rules may introduce subjective bias and may require recalibration for deployment across different student populations or institutional settings.

Seventh, the current study does not evaluate long-term deployment feasibility, real-time intervention effectiveness, or user engagement under continuous operational conditions. The prototype deployment interface demonstrates practical applicability; however, large-scale real-world validation remains necessary before institutional deployment can be considered.

Finally, ethical and privacy considerations remain important challenges in behavioral sensing research. Continuous monitoring of behavioral signals, psychometric responses, and digital activity patterns raises concerns related to data privacy, informed consent, secure storage, and responsible use of sensitive mental health information. Any future deployment of such systems would require rigorous ethical oversight, transparency, and privacy-preserving data management protocols.

Despite these limitations, the proposed framework demonstrates the feasibility of integrating multimodal behavioral sensing, temporal representation learning, graph-based modeling, clustering, and interpretable fuzzy inference for scalable student wellbeing assessment. The study therefore provides a foundation for future research involving larger populations, longitudinal validation, adaptive intervention systems, and privacy-aware behavioral health monitoring frameworks.

## 7 Conclusion

This study presented an interpretable multimodal framework for student mental health and wellbeing assessment using behavioral sensing, temporal representation learning, graph-based embeddings, clustering, and fuzzy inference. The proposed framework integrates heterogeneous data modalities including smartphone sensing signals, Ecological Momentary Assessment (EMA) responses, academic indicators, and psychometric survey data to model behavioral patterns associated with student wellbeing.

A bidirectional LSTM autoencoder was employed to learn latent temporal behavioral representations from sequential day-level sensing data, while graph-based representation learning captured structural similarities among students using similarity-driven neighborhood graphs. These multimodal representations were subsequently fused and analyzed using nonlinear dimensionality reduction and unsupervised clustering techniques to identify meaningful behavioral risk groups.

Experimental evaluation demonstrated that the proposed framework achieved meaningful clustering quality and stable behavioral stratification across low-risk, moderate-risk, and high-risk student profiles. The observed clusters exhibited interpretable differences in stress behavior, sleep consistency, social interaction, and academic engagement patterns, indicating that multi-modal behavioral modeling can effectively capture meaningful wellbeing-related characteristics.

To improve interpretability and practical usability, a hybrid fuzzy inference framework was additionally developed to compute Mental Risk Index, Academic Risk Index, and Overall Well-being Index using psychometric and behavioral representations. The integration of fuzzy reasoning enabled transparent and explainable wellbeing assessment while also supporting personalized recommendation generation for behavioral intervention guidance.

Compared with traditional statistical approaches and purely black-box predictive models, the proposed framework balances deep behavioral representation learning with interpretable reasoning and practical deployment capability. The inclusion of a prototype mobile application further demonstrates the feasibility of integrating the proposed framework into accessible educational wellbeing support systems.

Although the study is limited by dataset size, self-reported measurements, and lack of clinical validation, the findings demonstrate the potential of combining multimodal sensing, temporal learning, graph embeddings, clustering, and fuzzy inference for scalable behavioral wellbeing assessment in educational environments.

Future work will focus on extending the framework to larger and more diverse student populations, incorporating real-time adaptive intervention strategies, improving personalization through continual learning approaches, and exploring privacy-preserving behavioral modeling techniques. Additional research involving longitudinal clinical validation and real-world deployment studies may further strengthen the applicability of multimodal behavioral assessment systems for proactive student wellbeing support.

## Data Availability

All data processed in the present study are available upon reasonable request to the authors.

## 8 Ethics Statement

This study utilizes the publicly available StudentLife dataset collected and released by the original dataset authors under established research protocols [6]. All behavioral, sensing, academic, and psychometric data used in this work were anonymized prior to public release, and no personally identifiable information was accessed during the course of this study.

The proposed framework is intended solely for research-oriented behavioral wellbeing assessment and early risk identification. The system is not designed to function as a clinical diagnostic tool and should not be interpreted as a substitute for professional psychiatric evaluation, medical diagnosis, or therapeutic intervention.

Behavioral sensing and mental health modeling involve sensitive personal information, including psychological indicators, behavioral patterns, and digital activity data. Consequently, ethical considerations related to privacy, informed consent, secure data management, transparency, and responsible use of behavioral information are critically important for any future deployment of such systems.

The prototype deployment interface described in this study was developed exclusively for demonstrating the practical applicability of the proposed framework and was not deployed in a live institutional environment. No real-time behavioral monitoring or intervention was performed on human participants as part of this work.

Any future large-scale deployment of multimodal behavioral monitoring systems should incorporate rigorous ethical oversight, participant consent mechanisms, secure data storage protocols, privacy-preserving computation techniques, and clear institutional governance policies to ensure responsible and transparent use of sensitive wellbeing-related data.

## Code Availability

The code and implementation details associated with this study are available from the corresponding author upon reasonable request.

## Notes

### Competing Interest Statement

The authors have declared no competing interest.

### Funding Statement

his study did not receive any funding.

## References

[1] Fiona Wu, Ginger Freeman, Steve Wang, and Ingrid Flores. The future of college student mental health: Student perspectives. Journal of College Student Mental Health, 38(4):975–1010, 2024.

[2] Raquel Simões de Almeida, Andreia Rodrigues, Sofia Tavares, João F Barreto, António Marques, Maria João Trigueiro, Raquel Simões de Almeida, João Francisco Barreto, and António José Pereira da Silva Marques. Mental health and lifestyle factors among higher education students: a cross-sectional study. 2025.

[3] Sonya E Van Nuland, Victoria A Roach, Timothy D Wilson, and Daniel J Belliveau. Head to head: The role of academic competition in undergraduate anatomical education. Anatomical sciences education, 8(5):404–412, 2015.

[4] Maisam Ali, Shahid Ali, Qaiser Abbas, Zeeshan Abbas, and Seung Won Lee. Artificial intelligence for mental health: A narrative review of applications, challenges, and future directions in digital health. Digital Health, 11:20552076251395548, 2025.

[5] Steven Zhou, H Anne Weiss, Beth McCuskey, and Louis Tay. College student well-being: explaining academic and behavioral outcomes from a representative college student sample. Journal of Happiness Studies, 26(5):75, 2025.

[6] Rui Wang, Fanglin Chen, Zhenyu Chen, Tianxing Li, Gabriella Harari, Stefanie Tignor, Xia Zhou, Dror Ben-Zeev, and Andrew T Campbell. Studentlife: assessing mental health, academic performance and behavioral trends of college students using smartphones. In Proceedings of the 2014 ACM international joint conference on pervasive and ubiquitous computing, pages 3–14, 2014.

[7] Subigya Nepal, Wenjun Liu, Arvind Pillai, Weichen Wang, Vlado Vojdanovski, Jeremy F Huckins, Courtney Rogers, Meghan L Meyer, and Andrew T Campbell. Capturing the college experience: A four-year mobile sensing study of mental health, resilience and behavior of college students during the pandemic. Proceedings of the ACM on interactive, mobile, wearable and ubiquitous technologies, 8(1):1–37, 2024.

[8] Gordon Jackson-Koku. Beck depression inventory. Occupational medicine, 66(2):174–175, 2016.

[9] Duncan B Clark and John E Donovan. Reliability and validity of the hamilton anxiety rating scale in an adolescent sample. Journal of the American Academy of Child & Adolescent Psychiatry, 33(3):354–360, 1994.

[10] David Goldberg. Use of the general health questionnaire in clinical work. British medical journal (Clinical research ed.), 293(6556):1188, 1986.

[11] RE Lucas. Reevaluating the strengths and weaknesses of self-report measures of subjective well-being. Handbook of well-being, pages 1–12, 2018.

[12] Li Ruihua, Norlizah Che Hassan, Zhu Qiuxia, Ouyang Sha, and Dong Jingyi. A systematic review on the impact of social support on college students’ wellbeing and mental health. Plos one, 20(7):e0325212, 2025.

[13] Xiuyu Shi, Jin Pan, Daofu Yuan, Minye Li, and Yafeng Pan. Advanced data analysis and prediction model for student mental health risk assessment. Frontiers in Psychology, 16:1682083, 2025.

[14] Irene Bonafonte, Cristina Bustos, Abraham Larrazolo, Gilberto Lorenzo Martínez Luna, Adolfo Guzmán Arenas, Xavier Baró, Isaac Tourgeman, Mercedes Balcells, and Agata Lapedriza. Analyzing the contribution of different passively collected data to predict stress and depression. In 2023 11th International Conference on Affective Computing and Intelligent Interaction Workshops and Demos (ACIIW), pages 1–4. IEEE, 2023.

[15] Rui Wang. Mental health sensing using mobile phones. PhD thesis, 2018.

[16] Gatis Mikelsons, Matthew Smith, Abhinav Mehrotra, and Mirco Musolesi. Towards deep learning models for psychological state prediction using smartphone data: Challenges and opportunities. arXiv preprint arXiv:1711.06350, 2017.

[17] Abhinav Shaw, Natcha Simsiri, Iman Deznaby, Madalina Fiterau, and Tauhidur Rahaman. Personalized student stress prediction with deep multitask network. arXiv preprint arXiv:1906.11356, 2019.

[18] Enrique Delahoz-Domínguez, Adel Mendoza-Mendoza, and Delimiro Visbal-Cadavid. Clustering of countries through umap and k-means: A multidimensional analysis of development, governance, and logistics. Logistics, 9(3):108, 2025.

[19] Ke Liu, Jing Ma, and Edmund MK Lai. A dynamic fuzzy rule and attribute management framework for fuzzy inference systems in high-dimensional data. arXiv preprint arXiv:2504.19148, 2025.

[20] Weichen Wang, Shayan Mirjafari, Gabriella Harari, Dror Ben-Zeev, Rachel Brian, Tanzeem Choudhury, Marta Hauser, John Kane, Kizito Masaba, Subigya Nepal, et al. Social sensing: assessing social functioning of patients living with schizophrenia using mobile phone sensing. In Proceedings of the 2020 CHI conference on human factors in computing systems, pages 1–15, 2020.

[21] Byeongcheon Lee, Sangmin Kim, Jihoon Moon, Seungmin Rho, et al. Advancing autoencoder architectures for enhanced anomaly detection in multivariate industrial time series. Computers, Materials & Continua, 81(1), 2024.

[22] Asuka Tamaru, Junya Hara, Hiroshi Higashi, Yuichi Tanaka, and Antonio Ortega. Optimizing k in knn graphs with graph learning perspective. In ICASSP 2024-2024 IEEE International Conference on Acoustics, Speech and Signal Processing (ICASSP), pages 9441–9445. IEEE, 2024.

[23] Rosario Napoli, Gabriele Morabito, Antonio Celesti, Massimo Villari, and Maria Fazio. Improving graph embeddings in machine learning using knowledge completion with validation in a case study on covid-19 spread. arXiv preprint arXiv:2511.12071, 2025.

[24] Joshua B Tenenbaum, Vin de Silva, and John C Langford. A global geometric framework for nonlinear dimensionality reduction. science, 290(5500):2319–2323, 2000.

